# Diagnostic Accuracy of Vision-Language Models on Japanese Diagnostic Radiology, Nuclear Medicine, and Interventional Radiology Specialty Board Examinations

**DOI:** 10.1101/2024.05.31.24308072

**Authors:** Tatsushi Oura, Hiroyuki Tatekawa, Daisuke Horiuchi, Shu Matsushita, Hirotaka Takita, Natsuko Atsukawa, Yasuhito Mitsuyama, Atsushi Yoshida, Kazuki Murai, Rikako Tanaka, Taro Shimono, Akira Yamamoto, Yukio Miki, Daiju Ueda

## Abstract

**Purpose:** The performance of vision-language models (VLMs) with image interpretation capabilities, such as GPT-4 omni (GPT-4o), GPT-4 vision (GPT-4V), and Claude-3, has not been compared and remains unexplored in specialized radiological fields, including nuclear medicine and interventional radiology. This study aimed to evaluate and compare the diagnostic accuracy of various VLMs, including GPT-4 + GPT-4V, GPT-4o, Claude-3 Sonnet, and Claude-3 Opus, using Japanese diagnostic radiology, nuclear medicine, and interventional radiology (JDR, JNM, and JIR, respectively) board certification tests.

**Methods:** In total, 383 questions from the JDR test (358 images), 300 from the JNM test (92 images), and 322 from the JIR test (96 images) from 2019 to 2023 were consecutively collected. The accuracy rates of the GPT-4 + GPT-4V, GPT-4o, Claude-3 Sonnet, and Claude-3 Opus were calculated for all questions or questions with images. The accuracy rates of the VLMs were compared using McNemar’s test.

**Results:** GPT-4o demonstrated the highest accuracy rates across all evaluations with the JDR (all questions, 49%; questions with images, 48%), JNM (all questions, 64%; questions with images, 59%), and JIR tests (all questions, 43%; questions with images, 34%), followed by Claude-3 Opus with the JDR (all questions, 40%; questions with images, 38%), JNM (all questions, 51%; questions with images, 43%), and JIR tests (all questions, 40%; questions with images, 30%). For all questions, McNemar’s test showed that GPT-4o significantly outperformed the other VLMs (all *P* < 0.007), except for Claude-3 Opus in the JIR test. For questions with images, GPT-4o outperformed the other VLMs in the JDR and JNM tests (all *P* < 0.001), except Claude-3 Opus in the JNM test.

**Conclusion:** The GPT-4o had the highest success rates for questions with images and all questions from the JDR, JNM, and JIR board certification tests.

**Secondary abstract:** This study compared the diagnostic accuracy of vision-language models, including the GPT-4V, GPT-4o, and Claude-3, in Japanese radiological certification tests. GPT-4o demonstrated superior performance across diagnostic radiology, nuclear medicine, and interventional radiology tests, including image-based questions, highlighting its potential for medical image interpretation.

## Introduction

In recent years, the field of artificial intelligence (AI) has witnessed remarkable advancements, particularly in the development of large language models (LLMs) [1–4]. LLMs such as ChatGPT and Claude have demonstrated the ability to understand and generate human-like text across a wide range of domains, showing excellent performance in various medical fields [5,6]. Several studies have investigated the performance of LLMs in the field of radiology [7–10]. These studies revealed that LLMs exhibit high diagnostic accuracy not only in general radiological knowledge but also in specialized areas such as thoracic radiology, neuroradiology, and musculoskeletal radiology [11–14]. The recent emergence of LLMs with image interpretation capabilities such as GPT-4 with vision (GPT-4V) and Claude, which are often referred to as vision-language models (VLMs), has opened new possibilities for AI-assisted medical support. VLMs are designed to process and understand both visual and textual information, which enables them to analyze medical images and provide diagnostic insights. Among these models, the GPT-4 omni (GPT-4o), released by OpenAI in May 2024, has recently gained attention as a VLM that demonstrates high performance in multilingual support and image understanding.

Despite the growing interest in VLMs, only a few studies have evaluated their diagnostic accuracy in radiology quiz cases and specialty board examinations [15,16]. In particular, regarding GPT-4o, no reports have assessed the diagnostic accuracy in the field of radiology or compared its diagnostic performance among different VLMs. Furthermore, the diagnostic accuracy of VLMs in radiology subspecialties, including nuclear medicine (NM) and interventional radiology (IR), remains unknown. Diagnosing in these specialized fields requires the interpretation of various images and modalities related to diseases, making it crucial to evaluate the performance of VLMs. As the application of VLMs continues to expand in healthcare, it is essential to investigate how well these advanced models can handle the complexities and nuances of NM and IR as well as diagnostic radiology (DR) fields.

This study aimed to evaluate various VLMs, including GPT-4V, GPT-4o, Claude-3 Sonnet, and Claude-3 Opus, and compare their diagnostic accuracy in the Japanese diagnostic radiology (JDR), Japanese nuclear medicine (JNM), and Japanese interventional radiology (JIR) board certification tests. By examining the diagnostic accuracy of these questions, the extent to which VLMs can be performed in highly specialized fields, such as advanced DR, NM, and IR, can be assessed. Understanding the areas in which VLMs are proficient and those in which they are less effective can be useful when considering their future use in supporting medical care with VLMs.

## Materials and methods

### Data collection

The case vignettes were collected consecutively over a 5-year period as following sources: the 28th (August 23, 2019), 29th (August 28, 2020), 30th (August 20, 2021), 31st (August 26, 2022), and 32nd (August 25, 2023) JDR tests; the 16th (July 7, 2019), 17th (October 25, 2020), 18th (June 20, 2021), 19th (June 26, 2022), and 20th (July 2, 2023) JNM tests; and the 18th (November 17, 2019), 19th (November 8, 2020), 20th (November 14, 2021), 21st (November 13, 2022), and 22nd (November 12, 2023) JIR tests. The tests were downloaded from the official websites of the respective societies. Duplicate vignettes during the data collection period were excluded. A selection flowchart of the questions is shown in Figure 1.

**Fig. 1.**
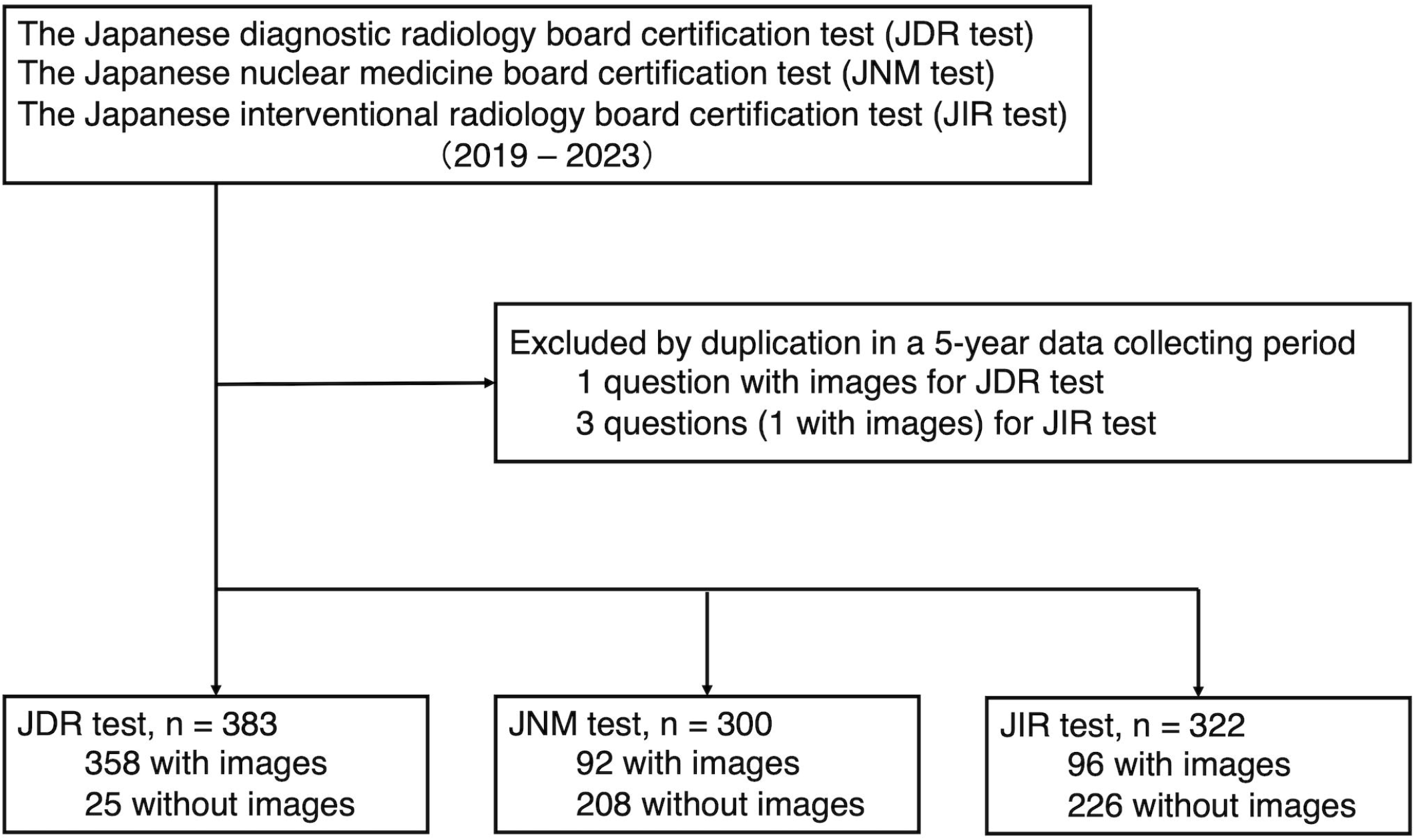
Selection flowchart of the questions JDR, Japanese diagnostic radiology; JNM, Japanese nuclear medicine; JIR, Japanese interventional radiology.

The protocol of this study was approved by the institutional review board. As this study only utilized the publicly available data, the requirement for informed consent was waived.

### Data assessment

Question prompts, patient histories, and images (if available) from each case vignette were provided as inputs to the language models. We initiated the input prompt for each case as follows: “As a highly experienced professor of radiology with 30 years of expertise, you assist in radiology cases. Your role is to analyze questions, patient histories, and imaging findings to determine correct answer(s),” in accordance with a previous study [12]. Subsequently, the text of the questions and options was input into the VLMs’ input field to generate responses. In addition, when images were available, they were input simultaneously.

Responses from all VLMs were collected between March 31, 2024, and May 22, 2024. The data collection approach differed for each language model, where GPT-4-based ChatGPT (version gpt-4-0125-preview) provided answers only for questions without images, and GPT-4V-based ChatGPT (version gpt-4-vision-preview) provided answers only for questions with images. In contrast, GPT-4o-based ChatGPT (version gpt-4o-2024-05-13), Claude-3 Sonnet, and Claude-3 Opus (accessed on 15th Mar 2024, https://claude.ai/) provided answers to all questions, regardless of image presence. With regard to the characteristics of the collected questions, the JNM test followed a format in which one correct answer was chosen from five options. In contrast, the JRD and JIR tests included questions where one correct answer was chosen from five options as well as questions where multiple correct answers (mainly two) were chosen from five options. For questions with multiple correct answers, only responses that perfectly matched all the correct answers were considered accurate. For all language models, each question was answered three times, and the most frequent answer was considered the final answer because of the limited repeatability and robustness of the current language models [13]. If the VLMs generated three different answers to a question, the answer was considered incorrect. Additionally, if the image of a question was determined to be in violation of the terms of service of the VLMs and no response was generated, the question was considered to be answered incorrectly.

As official answers were not available, two DR specialists (HT, a radiologist with 14 years of experience, and DH, a radiologist with 7 years of experience), two NM specialists (AY, a radiologist with 15 years of experience, and HT, a radiologist with 14 years of experience), and two IR specialists (HT, a radiologist with 14 years of experience, and KM, a radiologist with 9 years of experience) independently evaluated each case in their respective fields and provided their answers. If needed, they referred to textbooks and the literature to determine the correct answers. If the answers were in agreement, they were considered correct. In case of disagreement, a consensus was reached to determine the correct answer.

### Statistical analyses

The accuracy rates of GPT-4 + GPT-4V, GPT-4o, Claude-3 Sonnet, and Claude-3 Opus were calculated for all questions, questions with images, questions without images, single-answer questions, and multi-answer questions. Diagnostic accuracy rates were compared among the language models using Cochran’s *Q* test and post hoc McNemar’s test [17]. *P* values <0.05 were considered statistically significant. Statistical analyses were performed using Python version 3.11.8 (Python Software Foundation, Wilmington, DE, USA).

### Use of large language models

This manuscript was proofread with the assistance of ChatGPT (GPT-4o architecture; OpenAI, https://chat.openai.com/), and all outputs were confirmed by the authors.

## Results

A total of 383 questions from the JDR test (358 with images and 60 with multiple answers), 300 questions from the JNM test (92 with images), and 322 questions from the JIR test (96 with images and 177 with multiple answers) were included. One question from the JDR tests and three questions from the JIR tests were excluded because they were duplicated in the 5-year data collecting period.

The number of correct answers and accuracy rates for the JDR, JNM, and JIR tests are presented in Tables 1, 2, and 3, respectively. The GPT-4o-based ChatGPT demonstrated the highest accuracy across all tests (JDR: all questions, 49%; questions with images, 48%; JNM: all questions, 64%; questions with images, 59%; JIR: all questions, 43%; questions with images, 34%). Claude-3 Opus showed consistent performance across all evaluation categories (JDR test: all questions, 40%; questions with images, 38%; JNM: all questions, 51%; questions with images, 43%; JIR test: all questions, 40%; questions with images, 30%), ranking second in accuracy rate after GPT-4o. All performances, except for Claude-3 Sonnet and Opus in the JNM test, exhibited higher accuracy rates for non-image-based questions than for image-based questions. No responses were generated for one question in the JDR test and two questions in the JNM test for either GPT-4V or GPT-4o, while responses were generated from all questions for Claude-3 Sonnet and Claude-3 Opus.

**Table 1.** Correct answer rates in the Japanese diagnostic radiology board certification test.

|  | All questions | Question with<br>images | Question without<br>images | Single-answer<br>question | Multi-answer<br>question |
| --- | --- | --- | --- | --- | --- |
| No. of questions | 383 | 358 | 25 | 323 | 60 |
| GPT-4 + GPT-4V | 145 (38%) | 128 (36%) | 17 (68%) | 125 (39%) | 20 (33%) |
| GPT-4o | 188 (49%) | 172 (48%) | 16 (64%) | 169 (52%) | 19 (32%) |
| Claude-3 Sonnet | 120 (31%) | 110 (31%) | 10 (40%) | 105 (33%) | 15 (25%) |
| Claude-3 Opus | 152 (40%) | 135 (38%) | 17 (68%) | 126 (39%) | 26 (43%) |
| Cochran's $Q$ | 40.37 | 37.31 | 8.16 | 45.34 | 5.9 |
| $P$ value | <0.001* | <0.001* | 0.043* | 0.041* | 0.085 |

\*Statistically significant.

**Table 2.** Correct answer rates in the Japanese nuclear medicine board certification test.

|  | All questions | Question with images | Question without images |
| --- | --- | --- | --- |
| No. of questions | 300 | 92 | 208 |
| GPT-4 + GPT-4V | 149 (50%) | 35 (38%) | 114 (55%) |
| GPT-4o | 191 (64%) | 54 (59%) | 137 (66%) |
| Claude-3 Sonnet | 120 (40%) | 27 (29%) | 56 (27%) |
| Claude-3 Opus | 152 (51%) | 40 (43%) | 85 (41%) |
| Cochran's $Q$ | 87.85 | 18.68 | 75.71 |
| $P$ value | <0.001* | <0.001* | <0.001* |

\*Statistically significant.

**Table 3.** Correct answer rates in the Japanese interventional radiology board certification test.

|  | All questions | Question with<br>images | Question without<br>images | Single-answer<br>question | Multi-answer<br>question |
| --- | --- | --- | --- | --- | --- |
| No. of questions | 322 | 96 | 226 | 145 | 177 |
| GPT-4 + GPT-4V | 111 (34%) | 29 (30%) | 82 (36%) | 58 (40%) | 53 (30%) |
| GPT-4o | 138 (43%) | 33 (34%) | 105 (46%) | 78 (54%) | 60 (34%) |
| Claude-3 Sonnet | 98 (30%) | 27 (28%) | 71 (31%) | 52 (36%) | 46 (26%) |
| Claude-3 Opus | 130 (40%) | 29 (30%) | 101 (45%) | 66 (46%) | 64 (36%) |
| Cochran's $Q$ | 17.29 | 0.88 | 18.97 | 14.57 | 6.04 |
| $P$ value | <0.001* | 0.83 | <0.001* | 0.004 | 0.11 |

\*Statistically significant.

The results of the pairwise McNemar’s tests are shown in Table 4, where Cochran’s *Q* test showed statistically significant differences. For all questions, McNemar’s tests showed that GPT-4o-based ChatGPT significantly outperformed the other VLMs (all *P* < 0.007), except for Claude-3 Opus in the JIR test. For questions with images, the GPT-4o-based ChatGPT outperformed the other VLMs in the JDR and JNM tests (all *P* < 0.001), except for Claude-3 Opus in the NM test.

**Table 4.** *P* values of the pairwise McNemar’s test for each board certification test.

|  | All question |  |  | Questions with images |  |  | Questions without images |  |  |
| --- | --- | --- | --- | --- | --- | --- | --- | --- | --- |
|  | JDR test | JNM test | JIR test | JDR test | JNM test | JIR test | JDR test | JNM test | JIR test |
| 4+4V vs. Sonnet | 0.02* | <0.001* | 0.21 | 0.09 | 0.23 | – | 0.07 | <0.001* | 0.21 |
| 4+4V vs. Opus | 0.63 | 0.06 | 0.094 | 0.63 | 0.55 | – | 0.99 | 0.006* | 0.09 |
| 4+4V vs. 4o | <0.001* | <0.001* | 0.007* | <0.001* | <0.001* | – | 0.99 | 0.005* | 0.007* |
| Sonnet vs. Opus | <0.001* | <0.001* | 0.003* | 0.02* | 0.059 | – | 0.07 | 0.003* | 0.003* |
| Sonnet vs. 4o | <0.001* | <0.001* | <0.001* | <0.001* | <0.001* | – | 0.11 | <0.001* | <0.001* |
| Opus vs. 4o | <0.001* | <0.001* | 0.40 | <0.001* | 0.059 | – | 0.99 | <0.001* | 0.40 |

JDR, Japanese diagnostic radiology; JNM, Japanese nuclear medicine; JIR, Japanese interventional radiology.
\*Statistically significant.

## Discussion

This study evaluated the diagnostic accuracy of various VLMs, including GPT-4V, GPT-4o, Claude-3 Sonnet, and Claude-3 Opus, in the Japanese radiological board certification tests, including the JDR, JNM, and JIR tests. The results demonstrated that GPT-4o exhibited the highest accuracy rates across all tests, whereas Claude-3 Opus consistently ranked second. These findings highlight the potential of VLMs in supporting medical care in highly specialized fields, such as advanced DR, NM, and IR.

To the best of our knowledge, this is the first study to evaluate and compare the diagnostic accuracy of multiple VLMs, including GPT-4V, GPT-4o, Claude-3 Sonnet, and Claude-3 Opus, using the JDR, JNM, and JIR tests, and the results showed that the GPT-4o-based ChatGPT had the highest accuracy rates for questions with images as well as all questions. The superior performance of GPT-4o-based ChatGPT can be attributed to the broader and more recent dataset on which it was trained. Unlike Claude-3, which was trained only on data up to August 2023, GPT-4o-based ChatGPT incorporated data available up to December 2023. This extended training period likely provides more up-to-date information and advancements, contributing to higher diagnostic accuracy. In addition, GPT-4o not only has improved image recognition capabilities compared with GPT-4 but also shows remarkable enhancements in non-English languages, including Japanese. This could explain the better performance of the model using the GPT-4o in the evaluation of Japanese tests. By contrast, LLM drift, which refers to the deterioration in the performance of LLMs, may have influenced the performance of relatively old VLMs because of the impact of model updates and weight changes on maintaining the reliability of language models [13].

Given the varied difficulty levels of the questions, a simple comparison is not feasible. However, when comparing questions with images to questions without images, all VLMs demonstrated higher accuracy rates in questions without images, with the exception of Claude-3 Sonnet and Opus in the NM test. These results suggest that current VLMs have an inadequate capability to process radiological images and extract imaging findings. OpenAI, which developed ChatGPT, officially commented that the current GPT-4V is unsuitable for medical image interpretation and cannot replace professional medical diagnoses [18]. Furthermore, a previous study indicated that GPT-4V primarily depends on linguistic cues for decision-making with images supplementary [15]. Thus, future update and weight changes of VLMs in decision-making may vary the results. Techniques, such as retrieval-augmented generation, fine-tuning with reinforcement learning from appropriate feedback, and training vision models on a wide range of medical images, may also improve the performances [19]. Nonetheless, GPT-4o, the latest model of ChatGPT, showed the best performance in answering the test; therefore, VLMs are gradually improving their ability to recognize medical images.

In the JIR tests, although GPT-4o exhibited the highest accuracy rates among the other VLMs, the differences in performance between the models were less pronounced, and the overall accuracy was lower in the JIR test than in the other tests. This could be attributed to the trends in the JIR tests, which predominantly featured multiple-choice questions (JDR test, 60/383, 16%; JNM test, 0/300, 0%; JIR test, 177/322, 55%). Although a simple comparison between single– and multiple-answer questions might be inappropriate because of the varied difficulty levels of the questions, most VLMs demonstrated higher accuracy rates for single-answer questions. This trend suggests that language models may be better at handling questions in which only one correct answer needs to be identified, potentially owing to less complexity in the decision-making processes [20]. Additionally, JIR tests often require decisions not only for diagnosis but also for treatment options, which can vary based on the clinical scenario. This variability, along with questions demanding detailed anatomical knowledge, might have influenced the lower performance rates observed among the VLMs in this specialized field.

This study had several limitations. First, the questions used in this study may have been included in the training data of VLMs, which could introduce potential bias [21]. This bias may lead to overestimation of the diagnostic accuracy of VLMs [22]. Second, answering each question three times and using the most frequent response as the final answer may not be sufficient, as there could be variability with only three responses. This could lead to underestimation or overestimation of the performance of the VLMs. Third, this study evaluated the performance of Japanese questions. There may be differences in the performance of VLMs when using other languages, such as English. Fourth, because the official correct answers were not publicly available, there is a possibility that the answers provided by the specialists were incorrect, which might have prevented a fair evaluation of the performance of the LLMs.

In conclusion, this study evaluated the diagnostic accuracy of various VLMs in the JDR, JNR, and JIR board certification tests. The results demonstrated that GPT-4o exhibited the highest accuracy rates across all tests, whereas Claude-3 Opus consistently ranked second. The superior performance of GPT-4o can be attributed to its more recent and broader training dataset as well as its improved image recognition capabilities and enhancements in non-English languages. However, current VLMs have limitations in processing radiological images and extracting imaging findings. Despite these limitations, this study highlights the potential of VLMs to support medical care in highly specialized fields.

## Data Availability

The data that support the findings of this study are available on request to the corresponding authors.

## Acknowledgments

This study was supported by Guerbet and Iida Group Holdings.

## Notes

### Competing Interest Statement

The authors have declared no competing interest.

### Author Declarations

The study used (or will use) ONLY openly available human data that were originally located at: official websites of the Japan Radiological Society, Japanese Society of Nuclear Medicine, and Japanese Society Of Interventional Radiology

